# Facilitators and barriers to the implementation of a caregiver-led training programme for caregivers of children with cerebral palsy in rural Malawi

**DOI:** 10.1101/2025.10.22.25338418

**Authors:** TC Bakuwa, G Saloojee, W Slemming

## Abstract

**Background:** The implementation of caregiver-led training programmes for complex childhood-onset conditions like cerebral palsy is challenging as multiple factors influence outcomes, particularly in rural, resource-constrained settings. Understanding the perspectives of the caregivers and professionals implementing programmes is important to identify practical, cultural, and logistical factors that influence programme outcomes. This qualitative study explored the facilitators and barriers encountered during the implementation of a caregiver-led training programme in rural Mangochi, Malawi.

**Methods:** In-depth interviews were conducted with physiotherapists and caregivers of children with cerebral palsy who participated in a seven-week feasibility trial of the Malamulele Onward Carer-2-Carer Training programme in August 2023. Semi-structured interview guides informed by the Consolidated Framework for Implementation Research were used to conduct the interviews. The interviews were audio-recorded and transcribed verbatim. Data were managed using NVivo and analysed thematically. An abductive analytical approach was used, whereby themes were initially generated inductively and later interpreted deductively within the framework to enhance conceptual depth.

**Results:** Facilitators of programme implementation included the integration of training into daily caregiving routines, active caregiver engagement, and the logistical support provided by a community-based organisation. A key finding was the unique contribution of expert caregivers, who facilitated sessions using shared language, lived experience, and emotional connection, which enhanced caregiver participation and trust. The expert caregivers’ ability to mobilise local resources further supported programme delivery. Barriers included time constraints for covering some modules, slow learning and particular skill acquisition challenges among older caregivers and external factors such as poor access to transportation and limited family support for home practice. While both expert caregivers and therapists effectively facilitated the programme, expert caregivers encountered occasional challenges with content presentation when delivering sessions.

**Conclusion:** Reflections from caregivers and physiotherapists in rural Malawi highlighted the strengths of integrating lived experience into programme delivery. Expert caregivers fostered engagement and trust, helping to address participant learning challenges. However, continued mentorship, improving physical access and social support remain crucial for sustaining caregiver-led training programmes and maximising their impact on children with cerebral palsy and their families.

## INTRODUCTION

Cerebral palsy (CP) is the most common motor disability in children. Daily living is demanding for children and their families due to the lifelong activity and participation restrictions they face [1–3]. Caring for a child with CP in Low- and Middle-Income Countries (LMICs), such as rural Malawi, is particularly difficult, given the limited access to specialised healthcare, supportive services, and rehabilitation [1]. Caregivers, usually family members, are the primary providers of care and support for children with CP [4], a role and responsibility for which many feel unprepared. Caregiver strain, stress, fatigue, depression, and ultimately a reduced quality of life, are well-documented in the literature [1,5–8]. In addition, the high levels of poverty, stigma, and negative attitudes that caregivers living in LMICs face highlight both the pressures and the high levels of resilience that they need to demonstrate to sustain caregiving.

In response to these challenges, caregiver-led training programmes have been developed to bridge the rehabilitation service gap and to empower caregivers with the knowledge and skills needed to improve the quality of care for children with CP [9,10]. These programmes aim to leverage on the agency of the caregivers themselves as providers of peer support to enhance caregivers’ competencies in managing the daily needs of their children, promote the children’s physical and cognitive development, and alleviate some of the difficulties faced by caregivers [10–12]. While the benefits of caregiver-led training programmes have been described in literature, little is known about how they are implemented in resource-constrained settings. Understanding the factors that influence successful implementation will assist in ensuring sustainable integration of these programmes into resource-constrained health systems.

Effective implementation of caregiver-led training programmes depends on several factors. These include participants’ intrinsic motivation and attitude to peer support, which drives engagement and commitment [13], and sensitivity to the cultural and community needs of the setting [12,14,15]. Strong support networks further enhance the effectiveness and sustainability of these initiatives by providing resources and encouragement [10]. Additionally, active participation from community stakeholders fosters trust and collaboration, strengthening programme uptake [16]. While these factors are recognised as important, there is limited understanding of how they intersect and affect programme delivery in LMICs.

LMICs face significant challenges that potentially threaten programme implementation, including poor physical accessibility, socioeconomic constraints and shortage of time [17–19]. Moreover, structural issues within the training programmes, including insufficient tailoring to the local context and challenges in delivering complex content, often impede the achievement of intended learning outcomes [16,20,21]. Evidence from LMICs underscores the persistence of these challenges, which manifest in different patterns in various settings [1,17,21–24]. Addressing these issues requires a deep understanding of the specific contextual factors in these settings to inform the design of practical and sustainable solutions.

In Malawi, a feasibility trial of a caregiver-led training programme was conducted between January and August 2023, with a control group of caregivers trained by physiotherapists. Quantitative analyses showed similar outcomes in both groups, including improvements in caregiver knowledge, self-efficacy, and quality of life as well as child activity and participation [25]. It is helpful to understand the factors that facilitated success and those that posed challenges at different levels of implementation. Therefore, this study aimed to describe facilitators and barriers to the implementation of a caregiver-led training programme for caregivers of children with CP in a rural setting in Mangochi district, located in the southern region of Malawi.

## MATERIALS AND METHODS

### Study design

This was a qualitative study grounded in phenomenology, using in-depth interviews (IDI) to elicit perspectives from caregivers and physiotherapists.

### Study setting

The study was conducted in August 2023, within the premises of Tiyende Pamodzi Community-Based Organisation (CBO) in Mangochi district, Southern region of Malawi.

Tiyende Pamodzi is a grassroots non-profit organisation that sustains its activities through local income-generating projects and periodic donations from international supporters. The organisation provides essential services such as monthly individualised physiotherapy sessions, access to assistive devices, and nutritional support for children with disabilities. Collaborating closely with the Mangochi Government District Hospital and other rehabilitation partners, the centre ensures comprehensive medical and rehabilitation care for its registered children. By January 2023, the centre was serving over 400 children with physical impairments, predominantly those with cerebral palsy (CP). To manage these extensive needs, the organisation employs one physiotherapist who is responsible for covering 12 subsidiary centres within its service area. Between January and August 2023, 83 caregivers of children with cerebral palsy were involved in a feasibility trial of a skills training programme called the Malamulele Onward Carer to Carer Training Programme.

### The Malamulele Onward Carer-to-Carer Training Programme (MOC2CTP)

The Malamulele Onward Carer-to-Carer Training Programme (MOC2CTP) is a targeted caregiver-delivered skills training programme focused on teaching caregivers to understand what CP is, identify the different types of CP, learn to problem-solve positioning and handling skills related to each type of CP eating and drinking, play, communication and cerebral visual impairment. These aspects are embedded in blending therapy with activities of daily living and looking at CP management as a way of life. The main outcome of the programme is improvement in caregiver knowledge and skills related to child positioning, feeding, mobilisation, play and engagement, communication and vision. This is expected to improve child-level outcomes; including child-mobility, self-care, feeding, and social skills. The programme also aims to improve caregiver well-being and quality of life.

The MOC2CTP was developed in collaboration with caregivers and designed to be delivered by caregivers of children with CP (*expert caregivers*) to fellow caregivers of children with CP in resource-constrained settings. It comprises seven workshops, each lasting between 2 and 2.5 hours. Each session includes an ice-breaker activity based on themes of psychosocial support; information on a particular topic; a larger section of practical experiential demonstrations, as well as group discussions. Table 1 provides an outline of the content covered in the MOC2CTP.

**Table 1.**
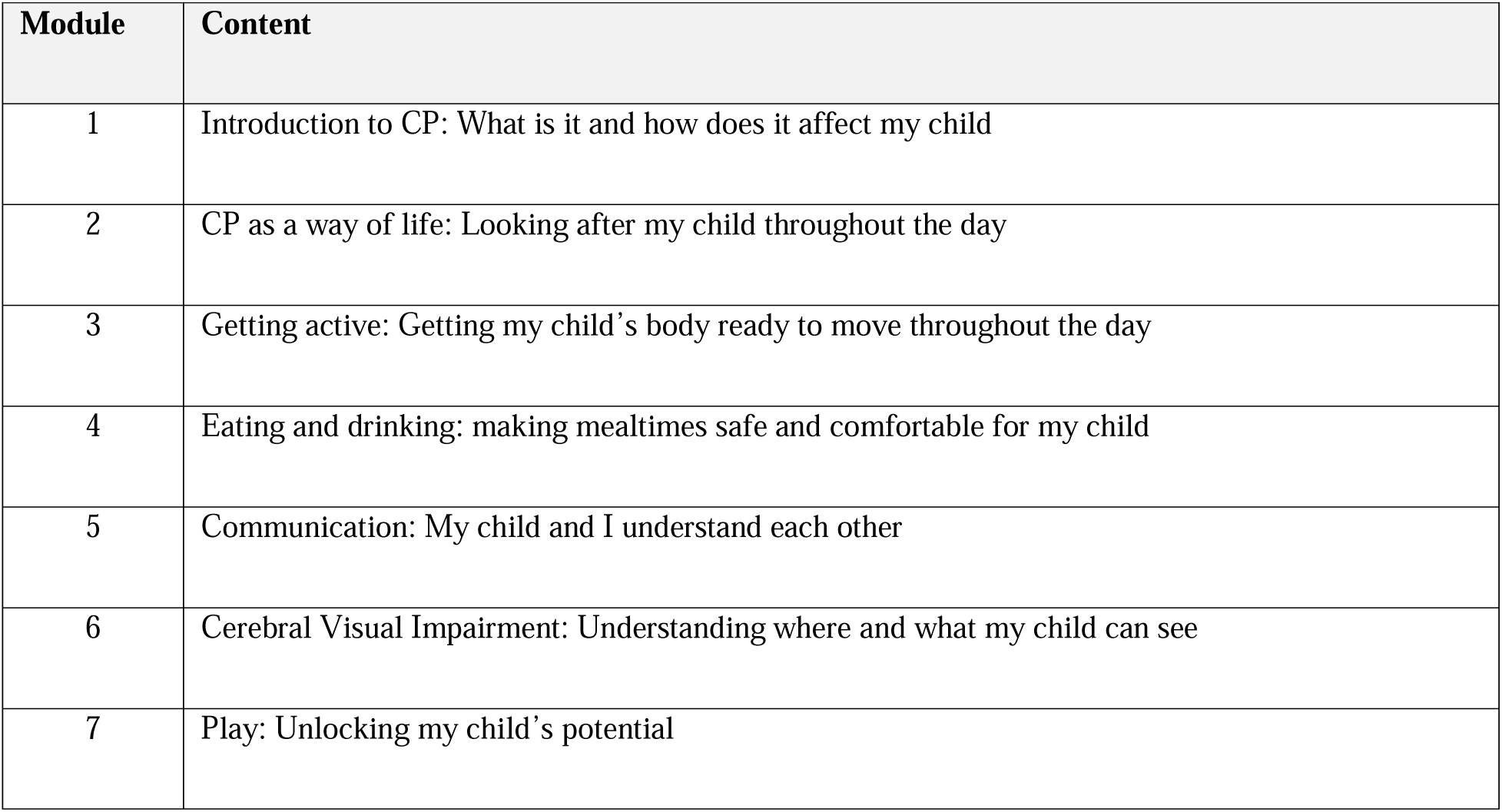
Content of the Malamulele Onward Carer-to-Carer programme.

### Implementation of the MOC2CTP in Malawi

The MOC2CTP was conducted in the form of a feasibility randomised controlled trial over seven weeks, with 83 caregivers randomly assigned in a 1:1 ratio to either the caregiver-led or therapist-led groups. Allocation was based on the Gross Motor Function Classification System (GMFCS), which categorises children with CP according to five mobility functioning levels: Level I being a child who can walk without limitations; Level II, a child who walks with some limitations but does not require assistive devices; Level III, a child who walks using a hand-held mobility aid; Level IV, a child who has limited self-mobility and may use powered mobility; and Level V, a child who is transported in a wheelchair and has severe limitations in head and trunk control [26]. Both groups received the MOC2CTP. The therapist-led arm was included as a comparison to assess whether caregiver-led training could achieve similar outcomes to conventional therapist-led approaches, which are often considered the standard in rehabilitation but may be less accessible in low-resource settings.

Caregivers attended workshops weekly for seven weeks, with each cohort of 10 caregivers completing one module per session. In the caregiver-led group, four expert caregivers working in pairs facilitated the sessions, while in the therapist-led group, two physiotherapists, each working on their own, conducted the sessions with the assistance of a trained translator. All sessions included psychosocial support activities, topic discussions, and hands-on practical demonstrations, utilising a picture-based guide and accessible materials such as feeding utensils, pillows, and balls.

Trainers (physiotherapists and expert caregivers) underwent extensive training before the programme’s implementation. The four expert caregivers received 30 hours of initial training from a master trainer, followed by additional workshops, supervised practice, and team facilitation training to enhance coordination. Both physiotherapists, one affiliated with the Tiyende Pamodzi CBO and the other from the district hospital, completed the same initial training and received additional material orientation either through workshops or remote consultation.

### Study participants

Study participants comprised all six trainers (four expert caregivers and two physiotherapists) and eleven caregivers who had attended the training programme as trainees. The caregivers were purposively selected to capture a broad range of experiences and perspectives, including variation in the child’s severity of cerebral palsy, as well as caregiver age and gender.

The two physiotherapists were in their early thirties and had over two years’ experience working with children with CP. The expert caregivers had prior experience supporting families of children with cerebral palsy and served as facilitators during the training sessions. Most of the caregivers were parents of children with cerebral palsy, while a few were grandparents or extended family members who assumed the primary caregiving role. Caregivers’ ages ranged from late teens to mid-forties, with the majority in their thirties. About half were married, while others were single, divorced, or widowed.

The ages of their children with cerebral palsy ranged from infancy to approximately nine years. Most of them had spastic cerebral palsy, while a smaller number had dyskinetic forms. Their functional ability as classified by the Gross Motor Function Classification System (GMFCS) varied from levels II to V.

### Data collection

Data were collected four weeks post-completion of the training programme. This four-week follow-up time was included to give time for caregivers to practice what they had learnt and provide feedback on the ease of implementation of the acquired information and skills in the home setting.

All in-depth interviews were conducted within the premises of the Tiyende Pamodzi CBO from the 26^th^ to the 30^th^ of August, 2023. The interviews were led by a female social worker with a Bachelor’s degree in Social Work (Community Development) and over seven years of experience in social work, counselling, and qualitative research within the region. She was supported by a female social work assistant, trained on the job and fluent in both local languages.

To enhance participant comfort and openness, particularly among male caregivers, two male Health Surveillance Assistants (HSAs) conducted interviews with the four male participants. Both HSAs had more than five years of experience working in community-based health programmes and collecting qualitative data. The involvement of interviewers who were not part of the programme implementation team was intentional, to minimise potential response bias and encourage open discussion of participants’ experiences. Participants were approached directly (face-to-face) and included in the study upon providing informed consent. Recruitment continued until no new information emerged from the interviews, with data saturation reached after interviewing the eleventh caregiver.

All interviews were audio-recorded with the consent of the study participants. Notes and observations during the interview were captured to aid triangulation and data analysis. Interviews with the caregivers who attended the MOC2CTP lasted an average of 40 minutes, while the interviews with the trainers (expert caregivers and physiotherapists) lasted an average of 65 minutes. Interview guides were informed by the Consolidated Framework for Implementation Research (CFIR), a useful tool for exploring implementation at multiple levels. It describes factors at various levels of implementation (viz programme delivery by facilitators/trainers and implementation at home by trainees/caregivers) which may affect the success of an intervention such as a training programme [27,28]. These factors include the design and features of the intervention itself, the people involved in delivering and receiving it, the immediate setting or environment in which the programme is delivered (such as clinics or community groups), broader external factors (such as community norms, policies, or available resources), and the overall process of implementation. The framework supports a comprehensive approach to identifying and addressing factors that influence key implementation outcomes, such as feasibility, acceptability, and sustainability [28]. Table 2 describes the CFIR constructs and how these were captured in the interview guides.

**Table 2.** Description of the CFIR constructs and related interview guide questions.

| CFIR Construct | Description from CFIR | Example of interview-guide questions based on the CFIR construct |
| --- | --- | --- |
| Intervention characteristics | All codes related to the attributes of the programme that made it easy/hard for the participants to implement it, including quality, complexity and relevance | What aspects of the programme did you find most useful or challenging?<br>How easy or difficult was it to implement the training activities?<br>**Prompt:** What made them easy/difficult? |
| Individual characteristics | All codes describing personal attributes of trainers or participants that made it easy/hard | What skills or personal attributes helped you implement the training/ what you learnt? |
|  | for them to implement the programme | What personal challenges did you face in implementing the programme/ the learnt skills?<br>**Prompt:** What previous experience was useful? |
| Inner context | All codes related to organisational factors that enabled/hindered implementation of the programme such as resource availability, leadership support and organisational support | What organisational support/barriers, if any, did you experience while implementing the programme/ while applying the skills at home?<br>**Prompt:** How supported did you feel, give examples |
| Outer context | All codes related to external factors that enabled/hindered implementation of the programme such as community access to resources, family needs and external policies | In what ways did your family or community influence your experience with the programme?<br>What factors in your community influenced your ability to participate in the programme?<br>**Prompt:** How did e.g. travel, family roles/ attitudes affect your participation |
| Process | All codes related to strategies for programme delivery and implementation that enhanced/hindered the implementation of the programme such as engagement styles, planning and delivery skills | What strategies were used to introduce the programme to participants?<br>- How were participants engaged throughout the implementation process?<br>- What planning strategies helped make the programme more effective?<br>**Prompt:** What communication styles worked/did not work, what could be done differently |

### Reflexivity

All three authors are qualified physiotherapists. TB (Msc) has over five years of experience working with children with cerebral palsy (CP), though this was her first engagement with the MOC2CTP. She served as the principal investigator for the study, focusing mainly on coordination of data collection and analysis. TB had no prior relationships with the caregivers or CBO representatives involved but had previously interacted with both physiotherapists during her undergraduate training and clinical internship.

WS and GS (PhDs) have extensive expertise in working with children with disabilities, including CP, and have significant research experience in this field. GS, who spearheaded the development of the MOC2CTP, has long been involved in assessing the programme’s applicability in various local and regional contexts.

The authors consistently reflected on their previous knowledge throughout the research process, ensuring that the analysis remained data-driven. Their collective experience significantly enhanced the depth of analysis and reflective inquiry.

### Data management and analysis

All audio-recorded interviews were transcribed verbatim in the original local languages, Yao and Chichewa, using Microsoft Word. Member checking was conducted by sharing transcripts with the physiotherapists and discussing selected transcripts with caregivers where possible, to verify the accuracy of the information and clarify any ambiguities.

The verified transcripts were subsequently translated into English and imported into NVivo version 12 (QSR International) for coding and analysis. Data were analysed thematically, guided by the Consolidated Framework for Implementation Research (CFIR) to explore facilitators and barriers across five levels: (i) intervention characteristics, (ii) individual characteristics, (iii) inner setting, (iv) outer setting, and (v) implementation process [13].

An abductive analytic approach was used, combining inductive and deductive reasoning [29]. TB initially conducted open, inductive coding to identify emerging, data-driven subthemes. WS and GS each coded one complete transcript independently to ensure code consistency and enhance interpretive credibility. All three authors then collaboratively reviewed and refined the coding framework, mapping subthemes deductively onto the CFIR constructs to deepen conceptual understanding.

Throughout the analytic process, the authors engaged in reflexive discussions to compare interpretations, minimise bias, and ensure that themes were grounded in participants’ accounts rather than researcher expectations. This iterative process supported the trustworthiness and confirmability of the findings.

## RESULTS

The findings are presented under two main categories reflecting the factors that facilitated or hindered the implementation of the caregiver-led training programme.

### Facilitators of programme implementation

The attributes that facilitated the implementation of the training programme were diverse and could be categorised under each of the five constructs of the CFIR. Three themes related to programme delivery related to the unique contribution that expert caregivers added: (i) shared language and experience which enhanced empathy, (ii) emotional connection and (iii) the ability to mobilise training resources from the community. The themes, subthemes and codes describing facilitators identified according to the CFIR framework are summarised in Table 3.

**Table 3.**
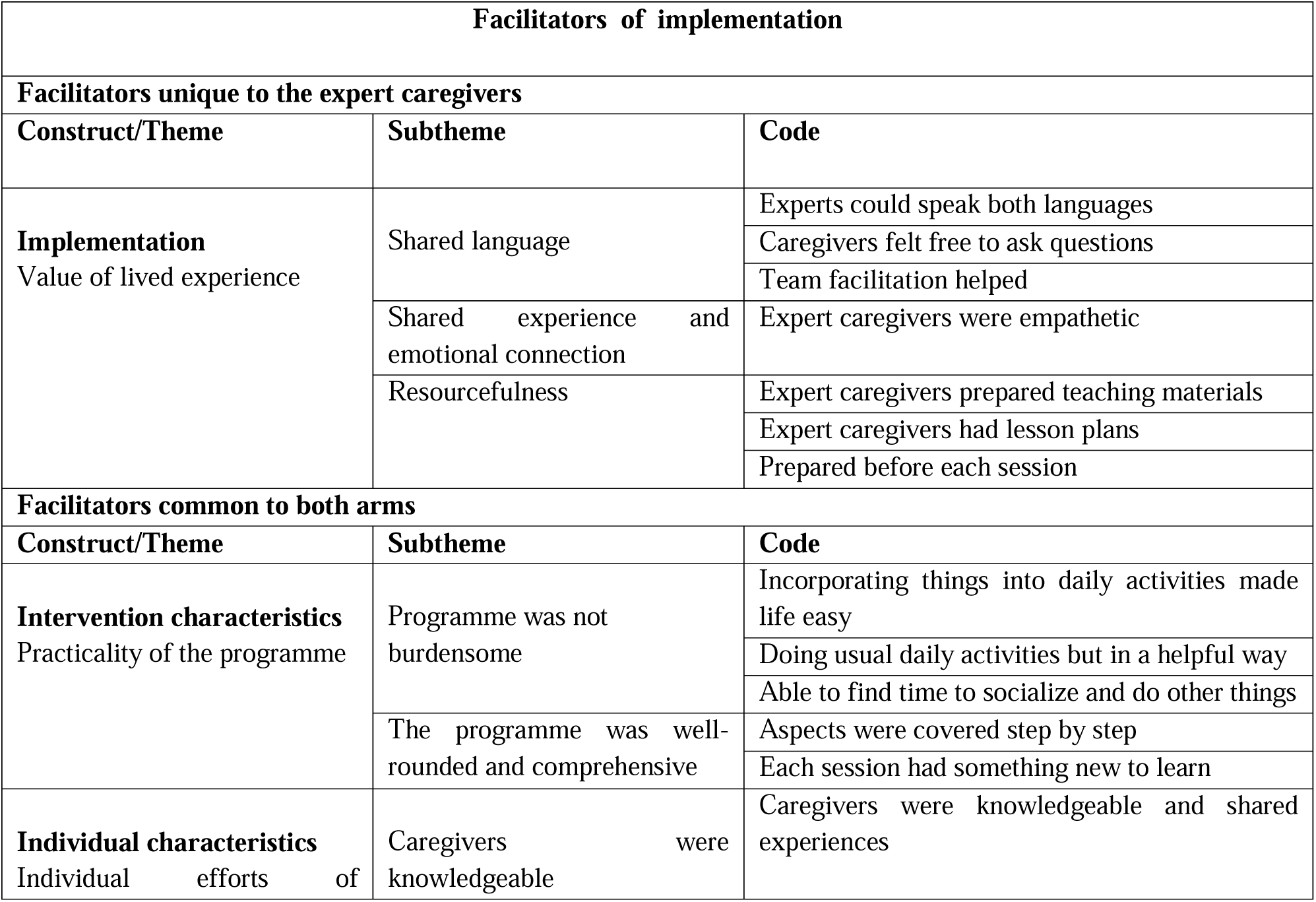
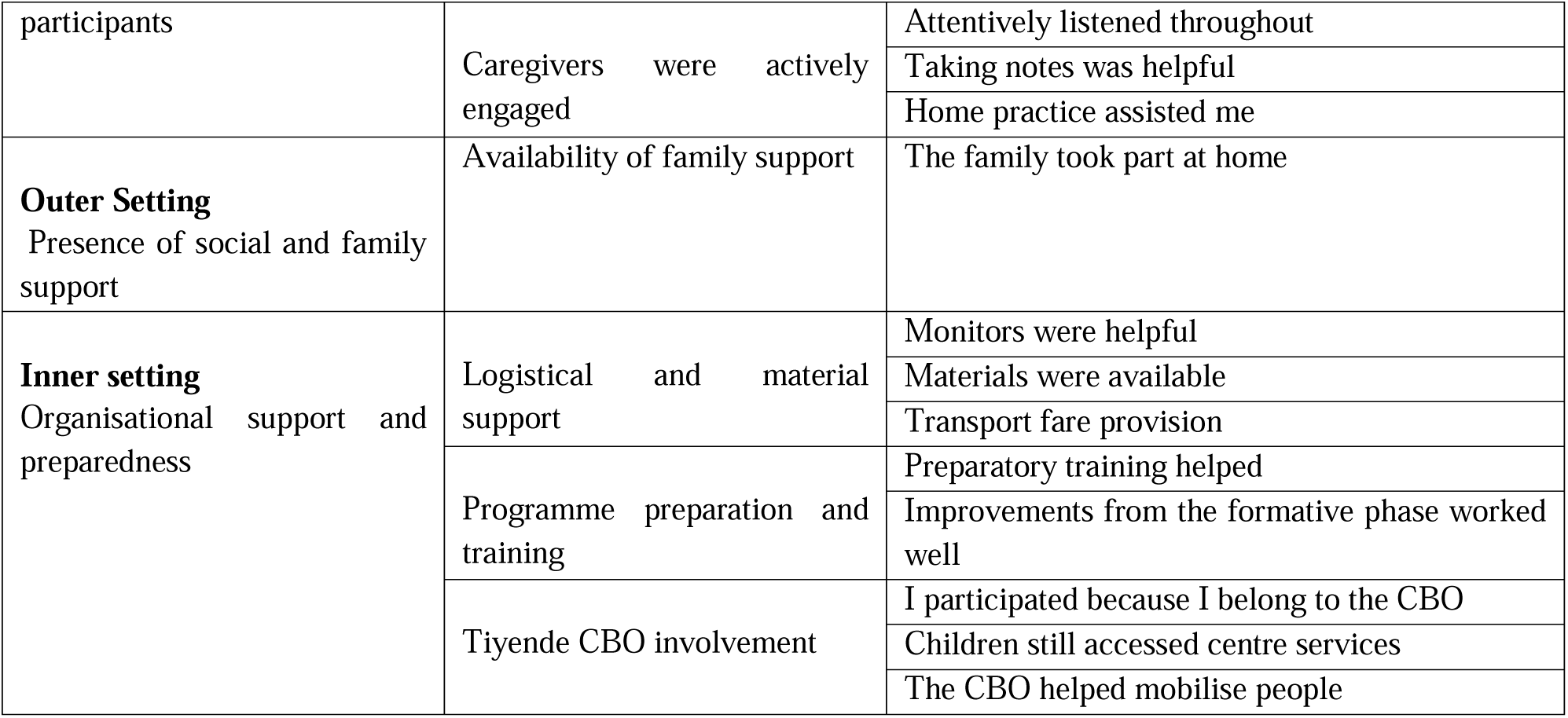
Facilitators related to implementation: themes, subthemes and codes.

| Facilitators of implementation |  |  |
| --- | --- | --- |
| Facilitators unique to the expert caregivers |  |  |
| Construct/Theme | Subtheme | Code |
| **Implementation**<br>Value of lived experience | Shared language | Experts could speak both languages |
|  |  | Caregivers felt free to ask questions |
|  |  | Team facilitation helped |
|  | Shared experience and emotional connection | Expert caregivers were empathetic |
|  | Resourcefulness | Expert caregivers prepared teaching materials |
|  |  | Expert caregivers had lesson plans |
|  |  | Prepared before each session |
| Facilitators common to both arms |  |  |
| Construct/Theme | Subtheme | Code |
| **Intervention characteristics**<br>Practicality of the programme | Programme was not burdensome | Incorporating things into daily activities made life easy |
|  |  | Doing usual daily activities but in a helpful way |
|  |  | Able to find time to socialize and do other things |
|  | The programme was well-rounded and comprehensive | Aspects were covered step by step |
|  |  | Each session had something new to learn |
| **Individual characteristics**<br>Individual efforts of | Caregivers were knowledgeable | Caregivers were knowledgeable and shared experiences |
| participants | Caregivers were actively engaged | Attentively listened throughout |
|  |  | Taking notes was helpful |
|  |  | Home practice assisted me |
| **Outer Setting**<br>Presence of social and family support | Availability of family support | The family took part at home |
| **Inner setting**<br>Organisational support and preparedness | Logistical and material support | Monitors were helpful |
|  |  | Materials were available |
|  |  | Transport fare provision |
|  | Programme preparation and training | Preparatory training helped |
|  |  | Improvements from the formative phase worked well |
|  | Tiyende CBO involvement | I participated because I belong to the CBO |
|  |  | Children still accessed centre services |
|  |  | The CBO helped mobilise people |

### Value of lived experience of expert caregivers

Expert caregivers’ lived experience enhanced programme delivery through **shared language**, **empathy**, and **resourcefulness**. Their ability to connect with fellow caregivers and organise training materials contributed to the programme’s effectiveness. The ability of expert caregivers to speak both local languages emerged as a key programme facilitator. While expert caregivers could speak in both languages, therapists needed a translator which delayed programme delivery.

> *“Even when others could not understand because of language, the trainers made sure to explain in both languages to make everyone understand the lessons. They were that good.”* (**Caregiver 5 caregiver-led arm**)
>
> *“Having the language interpreter was good. It helped me get through to those who only spoke Yao. Although the translation did make the session delivery longer than expected.”* (**Physiotherapist 2**)

Caregivers from both arms appreciated the kind reception by the trainers, irrespective of whether they were physiotherapists or expert caregivers. However, expert caregivers seemed to share an emotional connection with their fellow caregivers which was appreciated.

> *“We were also happy being taught by fellow women, because maybe, with the doctor some are afraid that they would not…. manage to ask because they were afraid. But our fellow woman, she understands us and she sees us.”* (**Caregiver 7 caregiver-led arm**)
>
> *“The therapist taught us well and was good to us. They made sure that we understood the type of the condition that our children have now we are able to differentiate that this is not a disease but a lifelong condition that we can live well with.”* (**Caregiver 8 therapist-led arm**)

The physiotherapists commended expert caregivers for their ability to independently organize and mobilize the necessary programme materials for both intervention arms, ensuring smooth programme delivery.

> *“These expert caregivers were really good. Each time I went to teach I would find everything I needed for facilitation already set, already in one place. They would have already prepared all the local material needed including for eating and drinking sessions! So, like after they had taught, they would pack everything and even set them up well for the next session. It made life easy for me.”* (**Physiotherapist 2**)
>
> *“Yes, even the rest of the volunteers who work there (Tiyende Pamodzi)were very supportive and so the project was going on well because of such people. Besides this we also had to go through a training before facilitating and that helped a lot. We were given the modules to use to teach including the materials which the expert caregivers prepared well.”* (**Physiotherapist 1**)

### Practicality of the MOC2CTP package

Participants found the training programme both practical and comprehensive, covering essential aspects of caregiving while remaining manageable within their daily routines. Caregivers played an active role in the learning process, demonstrating attentiveness and applying their prior knowledge to enhance their understanding which enhanced engagement. The programme covered various aspects of caregiving, including areas that caregivers had previously overlooked, such as play and communication with their children. The holistic approach of the MOC2CTP contributed to an enhanced understanding of childcare.

> *“I have learnt a lot about caring for my child than I expected. They touched all the areas of his daily care including things that I did not know were important like playing and talking to him.”* (**Caregiver 7, caregiver-led arm**)
>
> *“Ah mmm, as of now I can’t really think what we need to add because the way they made the modules, they cover many aspects, and they were going step by step…we learnt a lot that we did not know”* (**Caregiver 10, therapist-led arm**)

Despite their daily caregiving responsibilities, participants reported that integrating the content of the MOC2CTP training into their routines was manageable and practical. Many found that the content reinforced their existing activities, making it easier to implement therapy within everyday caregiving tasks.

> *“We do not have any worries, but now we take CP as a way of life and do physiotherapy on a daily basis because sometimes we lack time to do physiotherapy. So, whenever we are bathing, dressing, or standing the child, we need do it in helpful ways and to give toys too. This has helped me a lot.”* (**Caregiver 4, therapist-led arm**)
>
> *“I loved how the programme makes life easy. As a father I can help out now even know how to prepare his face for food. I wash his face and massage his face before feeding him in the morning. It does not demand much time.”* (**Caregiver 6, caregiver-led arm**)

One of the key strengths of the training experience was the high level of attentiveness demonstrated by caregivers receiving training. Their eagerness to learn contributed to engaged participation, allowing them to grasp and apply the lessons effectively.

> *“On the side of people being taught, there were no problems because they were attentive to see where the lessons would lead…and how they were going to apply them with their children.”* (**Expert Caregiver 3**)
>
> *“Among the parents we were teaching, the unity was good, they were eager to learn. It was seen in how we were able to ask each other questions and share”* (**Physiotherapist 1**)

The lived experiences of caregivers attending the workshops together with their prior knowledge of their children’s conditions enabled them to actively engage in discussions. Trainers were surprised by the depth of understanding participants already had, which made the learning process more interactive and meaningful.

> *“So, at first, maybe being adult learning, I would start with asking them questions, right? So, I was surprised the way they were responding, showing that they knew those things because they live with those children. Realising that oh, they know these things! It was really interesting.”* (**Physiotherapist 2**)
>
> *“I would also ask them if they have comments. There comments were, teacher what you are saying is true, because my child was failing this and that, but after starting physical exercises with him, now he is able. They had experiences to share”* (**Expert caregiver 1**)

### Organisational support and preparedness

The strong organisational support and trainer preparedness were key to the programme’s success. The community-based organisation (CBO) and research team ensured smooth delivery, while the trainers enhanced learning through practical, responsive engagement. The CBO and research team played an instrumental role in inviting caregivers and coordinating all activities. Their involvement ensured smooth implementation, access to resources, and consistent support for caregivers.

> *“Tiyende Pamodzi group has been so helpful, coming with my child here and finding this chance for therapies. And this programme also is just so helpful. But in other places, there is no organisation like Tiyende and no means to get programmes like these… So mine is still a plea that maybe, if there was a chance, communities should have organisations like these that can receive good programmes for people like me.”* (**Expert Caregiver 3**)
>
> *“I must say that the research team and Tiyende Pamodzi coordinated things well. The sessions ran smoothly, and we had all the equipment and resources that we needed. In addition to that, monitors were available to pick up any arising issues.”* (**Physiotherapist 1**)

All trainers (both expert caregivers and physiotherapists) demonstrated a high level of preparedness for programme delivery, which contributed to effective knowledge transfer. They engaged caregivers through practical demonstrations and revisions, ensuring that participants retained the skills taught.

> *“The therapist taught us well and was good to us. They made sure that we understood the type of condition that our children have, now we can differentiate that this is not a disease but a lifelong condition that we can live well with.”* (**Caregiver 8 therapist-led arm**)
>
> *“Our friends who were teaching had skills, they were well prepared. They demonstrated what they were teaching, and we could practice until we got it right. They showed us how to do things and kindly repeated where we missed and answered questions”* (**Caregiver 5 caregiver-led arm**)

### The presence of social and family support

Having family and social support was essential in enabling caregivers to attend training sessions. Some caregivers highlighted how their families took over caregiving duties, allowing them to focus on learning.

> *“I am grateful that my family accepted to take care of my child while I was here taking lessons because if they did not volunteer to help, I would not have managed.”* (**Caregiver 3 therapist-led arm**)
>
> *“Well since the programme I have not yet been to the center (referring to the subcentre where they receive therapy), since I was attending this programme. But my child went with my sister to the center, she helped me out.”* (**Caregiver 9, caregiver-led arm**)

### Barriers to programme implementation

Aspects that may have hindered implementation of the MOC2CTP were diverse and could be categorised under four of the five constructs of the CFIR. One theme, that of team coordination, highlights a challenge specific to programme delivery by expert caregivers. The themes, subthemes and codes describing barriers identified according to the CFIR framework are summarised in Table 4.

**Table 4.** Barriers to implementation: themes, subthemes and categories.

| Barriers to programme implementation |  |  |
| --- | --- | --- |
| Theme | Sub-theme | Codes |
| **Barrier unique to expert caregivers** |  |  |
| **Implementation process**<br>Expert caregiver delivery | Team coordination challenges | Training partner a bit behind |
| **General barriers common both arms** |  |  |
| **Intervention characteristic**<br>Complexity of some sections | Time constraints | Time not adequate to cover everything |
| **Individual characteristics**<br>Learner challenges | Learning and retention challenges | The elderly lost attention easily |
|  |  | Some were slow learners |
|  |  | Some needed more repeated practice |
| **Outer setting**<br>Absence of external support | Skill execution difficulties at home | The pain made mobilization difficult |
|  |  | Some techniques were difficult to perform at home |
|  | Lack of social or family support | No one to stay with my child |
|  | Transportation challenges | Hard to get motorcycles |

### Team coordination challenges

The effectiveness of the caregiver-led training was influenced by how well the two expert caregivers worked together as a team. Some expert caregivers struggled with coordination, particularly when one partner was less confident or behind in understanding the material. This occasionally disrupted the flow of training, leading to delays and the need for additional clarification as highlighted in the following:

> *“We did the training together, but my partner seemed behind a little bit. At times, she would not realise what page we were on. Yes… so sometimes it happened that maybe the pictures pasted are of a topic that’s ahead… So, sometimes we had to stop teaching and tell your friend to do this. That made us take longer than expected.”* (**Expert Caregiver 3**)

### Time constraints in some modules

The complexity of some sections of the MOC2CTP content required additional time for effective comprehension and skill acquisition by the caregivers. Both physiotherapists and expert caregivers highlighted the need for a slower pace, particularly when working with caregivers who had limited formal education or required more hands-on practice. The need for extended training time was seen as essential for ensuring a thorough understanding and retention of the material.

> *“Another thing, I see like for seven weeks to cover every lesson a day each, I see like the time is not enough, especially that we are teaching people who didn’t go to school. So, to take all the information at once, to grasp the whole lesson at once. It’s hard and so it needed time.”* (**Expert Caregiver 1**)
>
> *“I can say the time needs to be increased. Delivering the lessons in this setting needs a slower pace for the caregivers to grasp. We could use more time going forward.”* (**Physiotherapist 1**)

### Learning and retention challenges

Caregivers exhibited diverse learning abilities, with some being older, not having had formal education and requiring repeated practice to grasp new concepts effectively. This presented a challenge for all the trainers, who had to adapt their teaching strategies to accommodate the varied learning styles of the group while maintaining engagement and relevance for all.

> *“Mmm the most difficult, because maybe a good number of them are elderly and most of them completely did not go to school for them to have what we call attention.”* (**Physiotherapist 1**)
>
> *“Another thing, I see like for seven weeks to cover every lesson a day each, I see like the time is not enough, especially that we are teaching people who didn’t go to school. So, to take all the information at once, to grasp the whole lesson at once. It’s hard and so it needed time.”* (**Expert Caregiver 1**)

### Challenges applying some skills at home

Despite their enthusiasm for practicing the new ideas, some caregivers encountered challenges in applying certain interventions at home. Challenges included managing pre-existing pain in their children and difficulties in replicating some learned activities with the children at home.

> *“My child seems to be experiencing pain in his arm, he always cries even when I am moving it very gently like we were taught. He has always had this pain, perhaps because of the stiffness. I worry that I may hurt him.”* (**Caregiver 2, therapist-led arm**)
>
> *“Well, not difficult for me, but difficult for the child. Standing the child on the corner is difficult for him. He does not like it so much. And so, I have to really talk him into it. Some days it works, and sometimes it does not.”* (**Caregiver 6, caregiver-led arm**)

### Lack of social or family support

For some caregivers, limited social or family support made it difficult to ensure that the child remained engaged, supervised, and given freedom to interact and play as recommended during the programme. In the absence of assistance, some caregivers had to leave the child alone indoors while attending to other responsibilities.

> *“I have no family to leave the child with at home. And so, I still leave the child alone and lock her in the house. The child is quite big now. And so, it’s hard to say I should take her and go with her to do piece jobs. And so there I am still struggling.”* (**Caregiver 11, therapist-led arm**)

### Transportation challenges

Although there was provision for transport fare, some caregivers still encountered difficulties in accessing motorcycles or other transport options, especially those living in remote areas. These situations sometimes affected their ability to come on time or attend at all.

> *“Well, maybe I can talk about coming to this place. It was good that we were provided transport fare. However, some of us come from places that are far from the trading centre. Sometimes I had trouble getting a motorcycle and arriving on time. I think if the programme is brought to our sub-centres, things will be easier.”* (**Caregiver 7, caregiver-led arm**)

## DISCUSSION

This study aimed to describe factors that facilitated or hindered implementation of a caregiver-led training programme in rural Mangochi. Facilitators included the programme’s practical design, empathetic facilitators, a supportive learning environment, and strong logistical and social support. Barriers comprised time constraints for complex modules, limited formal education among some caregivers, transportation challenges, and insufficient social support for home practice. An important finding was the distinct contribution of expert caregivers, who enhanced programme delivery through their lived experiences.

By design, the Malamulele Onward Carer-to-Carer Training Programme (MOC2CTP) leverages lived experience as an educational tool, and the findings confirm its impact. It is not surprising that caregivers in the caregiver-led arm appreciated the emotional connection and empathy of expert caregivers during programme delivery. While similar programmes like the WHO Caregiver Skills Training (CST) are also emphasise programme delivery by non-specialists to improve training access [16], the MOC2CTP also highlights the unique role of peers with lived experience in bringing emotional and authentic relevance that enhances practical learning.

Despite their valuable contribution, the pairs of expert caregivers occasionally faced challenges in programme delivery, particularly when one caregiver was still struggling to articulate content from some parts of the programme manual. This underlines the need for continued mentoring, ongoing practice and structured team training to accommodate the different paces at which expert caregivers learn [16,30]. Furthermore, structured team training has been recognized as a key factor in improving facilitator confidence, promoting consistency in programme delivery, and ensuring long-term sustainability in community-based interventions [16,29]. Investing in these strategies could strengthen the capacity of expert caregivers and enhance the overall effectiveness of caregiver-led programmes.

Although the delivery of the MOC2CTP focuses on picture-based learning and practical demonstrations, the speed of learning among older participants and those who had little or no education was slower, highlighting another unique dimension of programme delivery in this setting [31,32]. Given that many caregivers in low- and middle-income countries (LMICs) may be older and have low literacy levels [32–35], it is essential to explore alternative methods to consolidate learning. Strategies such as increased practice time, more simplified visual aids, video-based demonstrations, or home-based reinforcement through home visits, may improve accessibility and retention among this population. This further adaptation of the materials and delivery strategies also entails continued further codesign with the caregivers to ensure relevant tailoring.

Participants found the application of the content covered in the MOC2CTP at home easy and manageable, aligning with evidence that user-friendly, adaptable interventions enhance adherence in low-resource settings [36]. However, social support played a crucial role in both programme participation and carryover of skills into the home setting. Caregivers with family support attended sessions consistently and implemented new skills more effectively, while those without support struggled to adopt caregiving practices. Additional challenges, such as managing impairments like pain, further limited application. These findings highlight the well-documented difficulty of translating learned skills into real-life settings [37,38] and emphasise the need for a supportive home environment. Addressing pain management within caregiver support interventions for caregivers of children with CP may therefore be critical. Strengthening peer support groups, remote supervision, and home visits could help sustain caregiver motivation and skill retention [37,38].

Transportation to the training venue emerged as a significant barrier to caregiver attendance, consistent with findings from rural health interventions in other global contexts [18,19,39]. Although transport fares were provided, some caregivers still struggled to access motorbike transport due to the remoteness of their homes, highlighting broader infrastructural and accessibility issues common in low-resource settings [39]. Addressing these transportation challenges, perhaps by decentralising programme delivery to the sub-centres as suggested, could improve caregiver attendance and extend the programme’s reach. Beyond this immediate context, these findings underscore a larger need for policymakers and government stakeholders to invest in rural transport systems and healthcare infrastructure. Such investment is essential to ensure that more families can access and benefit from community-based rehabilitation services.

### Strengths and limitations of the study

A key strength of this study is that it explored the perspectives of both caregivers and physiotherapists, offering a rich, contextualised understanding of implementation dynamics in a rural, resource-limited setting. The use of the Consolidated Framework for Implementation Research provided a robust theoretical lens for interpreting findings, while the abductive analytical approach allowed for both grounded insights and conceptual depth. Additionally, the inclusion of expert caregivers as central informants adds relevance to the study, given their dual role as implementers and community members.

However, a couple of limitations should be noted. First, the study drew on a small sample from a single seven-week feasibility trial conducted within one CBO in rural Mangochi. This narrow scope in both design and setting limits the transferability of findings to other contexts and larger-scale implementation. Secondly, social desirability bias may have influenced caregiver responses, particularly given their ongoing relationship with the CBO. Future studies should therefore test the programme across diverse settings, with larger samples and longer follow-up periods.

## CONCLUSION

The implementation of a caregiver-led training programme for children with CP in rural Mangochi, Malawi, demonstrated that such interventions are both feasible and valuable, particularly in resource-limited settings. Key facilitators included the programme’s non-burdensome design, its comprehensive and practical approach, caregivers’ active engagement and knowledge, skilled and empathetic facilitators, strong communication and support, and logistical assistance, including community mobilization by the Tiyende Pamodzi CBO. These strengths collectively enabled caregivers to acquire and apply new skills that directly supported their children’s care.

At the same time, several barriers were identified that may have hindered optimal implementation. These included limited time for delivering complex modules, learning and retention difficulties, delivery challenges among some expert-caregivers, lack of social-economic support, and transportation constraints. Addressing these issues through the provisions of more simplified learning materials, enhanced facilitator training, mentoring, structured follow-up, and stronger community-based support networks could further improve outcomes and sustainability. Importantly, the programme was effective despite these contextual limitations, underscoring its adaptability and potential scalability. The bottom line is that caregiver-led training, when embedded within trusted community structures and supported by both professional and peer facilitators, offers a practical, scalable, and sustainable strategy to strengthen care for children with CP in low-resource settings.

## Data Availability

All the data from which the conclusions have been made are included within the manuscript. A codebook for the qualitative data has been made available as a supplementary file (S1).

https://doi.org/10.5281/zenodo.17413770

## Data Availability

https://doi.org/10.5281/zenodo.17413770

## ETHICS APPROVAL AND CONSENT TO PARTICIPATE

Ethical approval was obtained from the Human Research Ethics Committee (Medical) at the University of Witwatersrand in South Africa (M220924) and the College of Medicine Research Ethics Committee at the Kamuzu University of Health Sciences in Malawi (P.04/22/3608). The purpose of the study was explained to all participants, including the researchers’ intention to use their reflections to inform future implementation of the programme. Participation was entirely voluntary, and written informed consent was obtained from all participants. For those unable to write, consent was documented using a thumbprint.

## AVAILABILITY OF DATA AND MATERIALS

All the data from which the conclusions have been made are included within the manuscript. A codebook for the qualitative data is available on Zenodo (https://doi.org/10.5281/zenodo.17413770).

## AUTHOR CONTRIBUTIONS

TB, WS, and GS contributed to the conception and design of the study. TB organised and coordinated the data collection. TB conducted the data analysis with the guidance of WS and GS. TB wrote the first draft of the manuscript and WS and GS reviewed the manuscript to the last draft. All authors contributed to the article and approved the submitted version.

## ACKNOWLEDGEMENTS

We would like to express our sincere gratitude to the participants for their time and dedication. Special thanks to the Tiyende Pamodzi CBO for hosting the study and seamlessly mobilising caregivers for the programme. We also thank the Malamulele Onward Organisation for offering training and support with MOC2CTP implementation. We appreciate the Mangochi District Hospital rehabilitation department for generously supporting the training of caregivers and evaluation of the programme.

